# Frontal subcortical executive dysfunction and minor hallucinations in Parkinson’s disease are linked to sensitivity to somatomotor conflicts

**DOI:** 10.1101/2025.08.22.25334259

**Authors:** Jevita Potheegadoo, Léa F. Duong Phan Thanh, Fosco Bernasconi, Nathalie H. Meyer, Laurent Jenni, Marie E. Maradan-Gachet, Cyrille Stucker, Sabina Catalano Chiuvé, Julien F. Bally, Mayte Castro Jimenez, Vanessa Fleury, Judit Horvath, Benoît Wicki, Javier Pagonabarraga Mora, Paul Krack, Olaf Blanke

**Affiliations:** Laboratory of Cognitive Neuroscience, Neuro-X Institute, School of Life Sciences, École Polytechnique Fédérale de Lausanne, EPFL, Geneva, Switzerland; Department of Neurology, Inselspital, University Hospital Bern, University of Bern, Bern, Switzerland; Graduate School for Health Sciences, University of Bern, Bern, Switzerland; Department of Neurology, Geneva University Hospital, Geneva, Switzerland; Department of Clinical Neurosciences, Service of Neurology, Lausanne University Hospital and University of Lausanne, Lausanne, Switzerland; Faculty of medecine, University of Geneva, CMU, Geneva, Switzerland; Neurology Center, La Tour Hospital, Meyrin, Switzerland; Neurocentre, Hôpital du Valais, Sion, Switzerland; Movement Disorder Unit, Neurology Department, Hospital de la Santa Creu i Sant Pau, Barcelona, Department of Medicine, Autonomous University of Barcelona, Spain; Department of Clinical Neurosciences, University Hospital Geneva, Geneva, Switzerland

**Keywords:** Parkinson’s disease, minor hallucinations, neuropsychology, frontal subcortical dysfunctions, subjective cognitive impairment

## Abstract

**Background:** Minor hallucinations (MH) affect 30-60% of patients with Parkinson’s disease (PD), and are considered precursors to structured visual hallucinations and cognitive decline. While the link between structured visual hallucinations and dementia is well established, the neuropsychological correlates of MH in PD remain unclear; most studies finding no significant cognitive differences between patients with MH and those without any hallucinations.

**Objectives:** Presence hallucinations (PH) being among the most prevalent MH in PD, we used a robotic procedure delivering somatomotor conflicts inducing PH experimentally to investigate whether sensitivity to such robot-induced PH aids in detecting cognitive differences between patients with MH and without hallucinations.

**Methods:** 31 PD patients with MH (PD-MH) and 37 without hallucinations (PD-nH) underwent neuropsychological assessment and the robotic procedure inducing PH. The sensitivity to report robot-induced PH was analyzed in relation to cognitive performance in neuropsychological tests.

**Results:** PD-MH patients reported more robot-induced PH than PD-nH patients, supporting previous findings. While both groups showed comparable performance in neuropsychological testing, we found a significant association between increased sensitivity to the PH-induction and poorer performance in frontal subcortical functions (executive functions), in PD-MH patients, but not in PD-nH patients.

**Conclusions:** These findings demonstrate that sensitivity to robot-induced PH reveals a previously undetected link between MH and frontal subcortical cognitive deficits in PD, pointing to shared underlying mechanisms between executive dysfunction and somatomotor processes involved in MH. This approach offers a novel and clinically valuable means of identifying early cognitive vulnerability that assessments relying only on standard testing may overlook.

**Plain language summary:** *Induced minor hallucinations are linked to executive function alteration in Parkinson’s disease:* This study investigated two groups of Parkinson’s disease (PD) patients: one group with minor hallucinations (MH) and another without any hallucinations. Using cognitive tests and a robotic task designed to temporarily induce a presence hallucination (hallucination of someone being there when no one is actually present), the study examined the relationship between cognitive impairment and sensitivity to robot-induced presence hallucinations (riPH). While both groups performed similarly on traditional cognitive tests, patients with MH were more sensitive to the riPH procedure. This increased riPH sensitivity was linked to difficulties with cognitive functions, especially executive functions, which are generally supported by the brain’s frontal and deeper regions (frontal subcortical network). These results from both tests (riPH, neuropsychology) suggest that elevated sensitivity to riPH is associated with mild signs of cognitive decline in PD patients that traditional tests might not detect. The use of the robotic induction of hallucinations as a tool for assessing cognitive deficits could offer a more sensitive method for identifying cognitive issues earlier in PD, potentially enabling earlier interventions.

## Introduction

Hallucinations are common and frequent non-motor symptoms of Parkinson’s disease (PD), affecting 30-60% of patients [1–3]. Hallucinations in PD can manifest across different sensory modalities and vary in their phenomenological complexity [4, 5]. Typically, structured or well-formed hallucinations, which are primarily visual in PD (e.g., seeing people, animals, objects), emerge in the mid to advanced stages of the disease [6]. However, many PD patients also experience so called “minor hallucinations” (MH) that occur frequently and earlier in the course of the illness, and precede structured visual hallucinations by many years and later co-occur with them [4, 6–8]. MH comprise presence hallucinations (PH) (sensation that someone is nearby despite no one being there), passage hallucinations (feeling of an indefinite figure passing in peripheral vision) and illusions (misperception of actual stimuli) [4]. Both structured hallucinations and MH can cause significant distress, negatively affecting patients’ quality of life, leading to increased caregiver burden, and complicating disease management with poor functional outcome [9–12]. Moreover, several clinical and research findings highlighted that hallucinations represent a risk factor for accelerated cognitive decline and dementia in PD, suggesting a more severe or rapidly progressing form of the disease [13–15].

Cognitive deficits in PD can be viewed as a spectrum ranging from subtle cognitive changes and fluctuations to mild cognitive impairment and ultimately, dementia [14, 16–18]. Generally, PD patients can show deficits across multiple cognitive domains, including attention, visual and verbal memory, visuospatial abilities, and executive functions [19–22]. Neuropsychological studies have consistently linked cognitive impairment and dementia to structured visual hallucinations, typical of advanced PD [8, 13, 23–27]. Non-demented patients with structured visual hallucinations often show mild cognitive impairment with deficits in frontal subcortical functions such as executive functions, attention and visuospatial abilities [28–32]. Research also indicated a sharp decline in posterior cortical functions (visual and spatial processing, language) during the transition to dementia in patients with structured visual hallucinations [8, 27, 30].

While MH are considered precursors to structured visual hallucinations and therefore a potential early marker of dementia [4, 33], MH have not been linked to global cognitive decline nor dysfunction in specific cognitive domains using neuropsychological tests, despite evidence of structural and functional brain changes in PD patients with MH [34–36]. Though a few studies suggested impairments of attentional control and visuospatial abilities in PD patients with MH compared to patients without hallucination [37, 38], the large majority of studies did not find any significant differences in global or specific cognitive functions between PD patients with MH compared to patients without hallucination, with both groups having similar neuropsychological performance (32–34).

We recently described a method to experimentally induce MH, particularly PH, using robotically-controlled somatomotor stimulation (robot-induced PH, riPH) [39]. This transitory controlled state of riPH has been studied in healthy and clinical populations, including PD [36, 40–43], and found that PD patients with MH in daily life reported stronger riPH (i.e., were more sensitive) compared to PD patients without any hallucinations [36]. Here, we examined cognitive function and riPH in a group of PD patients with MH (PD-MH) and without any hallucination (PD-nH). Based on our previous findings [36], we hypothesized that our PD-MH group would exhibit heightened sensitivity to riPH compared to the PD-nH group. Similar to previous work [7, 34, 38, 44], we did not expect significant differences in overall neuropsychological performance between the two groups as measured by a comprehensive neuropsychological test battery. However, because riPH activate frontal brain regions (inferior frontal gyrus and premotor cortex) [36], we explored whether, in PD-MH patients, greater sensitivity to riPH would be associated with poorer frontal subcortical functioning (e.g., executive functions including working memory, sustained attention, resistance to interference, mental flexibility, inhibitory control) [37, 38]. If confirmed, this would not only support the idea that the occurrence of MH in PD reflects frontal vulnerability, as hypothesized in previous work [7, 37, 38, 45], but also highlight the value of combining the riPH measure with neuropsychological testing that standard tests do not capture.

## Methods

### Participants and clinical assessment

Sixty-eight outpatients (21 women) diagnosed with PD based on the Movement Disorder Society (MDS) diagnostic criteria for PD [46], were included in the study. A semi-structured clinical interview was conducted to assess the occurrence and types of hallucinations in all patients. Those who experienced at least one MH recurrently, since disease onset and/or within the past month prior inclusion to the study, were classified as the PD-MH group (n=31), while those without any hallucination were categorized as the PD-nH group (n=37). The recruitment of patients was part of a larger multicenter project investigating hallucinations in PD. For the present study, given the primary focus on MH, patients experiencing structured visual hallucinations in addition to MH were not included. Socio-demographic and clinical data, including age, sex, disease duration, and levodopa equivalent daily dose (LEDD) [47, 48], were recorded. Severity of motor and non-motor symptoms of PD and stage of illness were evaluated using MDS-UPDRS and Hoehn and Yahr scales, respectively [49]. State of anxiety and depression were assessed with the Hospital Anxiety and Depression scale [50]. **Table 1** summarizes the demographic and clinical characteristics of patients.

**Table 1.** Socio-demographic and clinical characteristics of patients.

|  | PD-MH (n=31) |  | PD-nH (n=37) |  | Welch's t-test |  |  |
| --- | --- | --- | --- | --- | --- | --- | --- |
|  | M | SD | M | SD | t | df | P |
| **Age (years)** | 66.31 | 8.82 | 64.64 | 8.05 | 0.78 | 62.52 | 0.44 |
| **Education (years)** | 16.19 | 4.55 | 15.14 | 3.15 | 1.04 | 52.62 | 0.30 |
| **Illness duration (years)** | 10.62 | 4.31 | 9.81 | 4.68 | 0.88 | 65.23 | 0.38 |
| **Total LEDD (mg)** | 964.43 | 602.10 | 857.12 | 535.38 | 0.76 | 58.68 | 0.45 |
| **MDS-UPDRS I** | 14.52 | 7.02 | 8.83 | 5.86 | 3.55 | 59.25 | <0.001*** |
| **MDS-UPDRS II** | 13.00 | 6.83 | 10.44 | 6.29 | 1.51 | 62.78 | 0.13 |
| **MDS-UPDRS III** | 19.71 | 9.33 | 19.58 | 11.34 | 0.05 | 62.96 | 0.96 |
| **MDS-UPDRS IV** | 4.69 | 3.91 | 4.17 | 5.01 | 0.51 | 65.97 | 0.61 |
| **Hoehn & Yahr stage** | 1.93 | 0.74 | 1.85 | 0.58 | 0.50 | 52.48 | 0.62 |
| **Anxiety (HADS)** | 7.41 | 3.20 | 5.28 | 2.87 | 2.29 | 60.69 | 0.03* |
| **Depression (HADS)** | 6.28 | 3.85 | 4.08 | 2.82 | 2.12 | 54.71 | 0.04* |

\* $P < 0.05$ .
\*\* $P < 0.01$ .
\*\*\* $P < 0.001$ .
Abbreviations: LEDD, levodopa equivalent daily dosage; MDS-UPDRS, Movement Disorder Society- Unified Parkinson's Disease Rating Scale; HADS, Hospital Anxiety and Depression Scale.

Patients were not diagnosed with other neurological, psychiatric, or substance abuse disorders. Written informed consent was obtained from all patients, and the study was approved by Swiss cantonal ethics committees (protocol reference 2019-02275, 2021-01608), in accordance with the Declaration of Helsinki.

### Neuropsychological assessment

All patients were screened for cognitive impairments with the Montreal Cognitive Assessment (MoCA) [51], followed by a comprehensive assessment of major cognitive domains such as executive functions, attention, visuospatial abilities, memory and language using the Parkinson’s Disease Cognitive Rating Scale (PD-CRS) [30]. The PD-CRS consists of cognitive tests that separately assess frontal-subcortical dysfunctions (frontal subcortical score) and temporal posterior cortical impairment (posterior cortical score). We also tested patients’ performance in two other comprehensive neuropsychological tests on executive function: the Trail Making Test (TMT part A and B) (cognitive flexibility) [52] and the Stroop color-word test (resistance to interference, inhibitory control and cognitive flexibility) [53, 54] (see Supplemental Material 1). Finally, we assessed subjective cognitive impairment (SCI) of patients in daily life by means of a short interview and question 1.1 of MDS-UPDRS (“over the past week have you had problems remembering things, following conversations, paying attention, thinking clearly, or finding your way around the house or in town?”).

### Somatomotor stimulation task inducing PH

The experimental paradigm involving the robotic system was performed as described in previous work [36, 39, 40, 55]. A commercial haptic interface (Phantom Omni, SensAble Technologies) was placed in front of the patients, and integrated with a custom-made three-degree-of-freedom robot positioned behind the patient [39, 40]. Patients were seated and equipped with a blindfold and headphones delivering constant white noise to isolate them from external distractions. Participants were instructed to perform back-and-forth movements using the robotic device placed in front of them, with the hand they felt most comfortable using. The hand movement performed by patients was reproduced by the other robotic device located behind them, delivering tactile feedback on the participants’ back (see **Figure 1**). Participants were asked to perform the somatomotor task, while a pseudo-randomized delay of 0, 250 or 500 ms was introduced between the hand movement and the tactile feedback on the back. On each trial, each patient performed 10 successive poking movements (automatically counted) and immediately afterwards gave a “Yes” or “No” answer to the following question assessing riPH: “Did you feel as if someone was standing close by — behind or next to you?”. Each patient underwent 12 trials per delay (total of 36 trials). Breaks between sessions allowed patients to avoid physical discomfort and fatigue.

**Figure 1.**
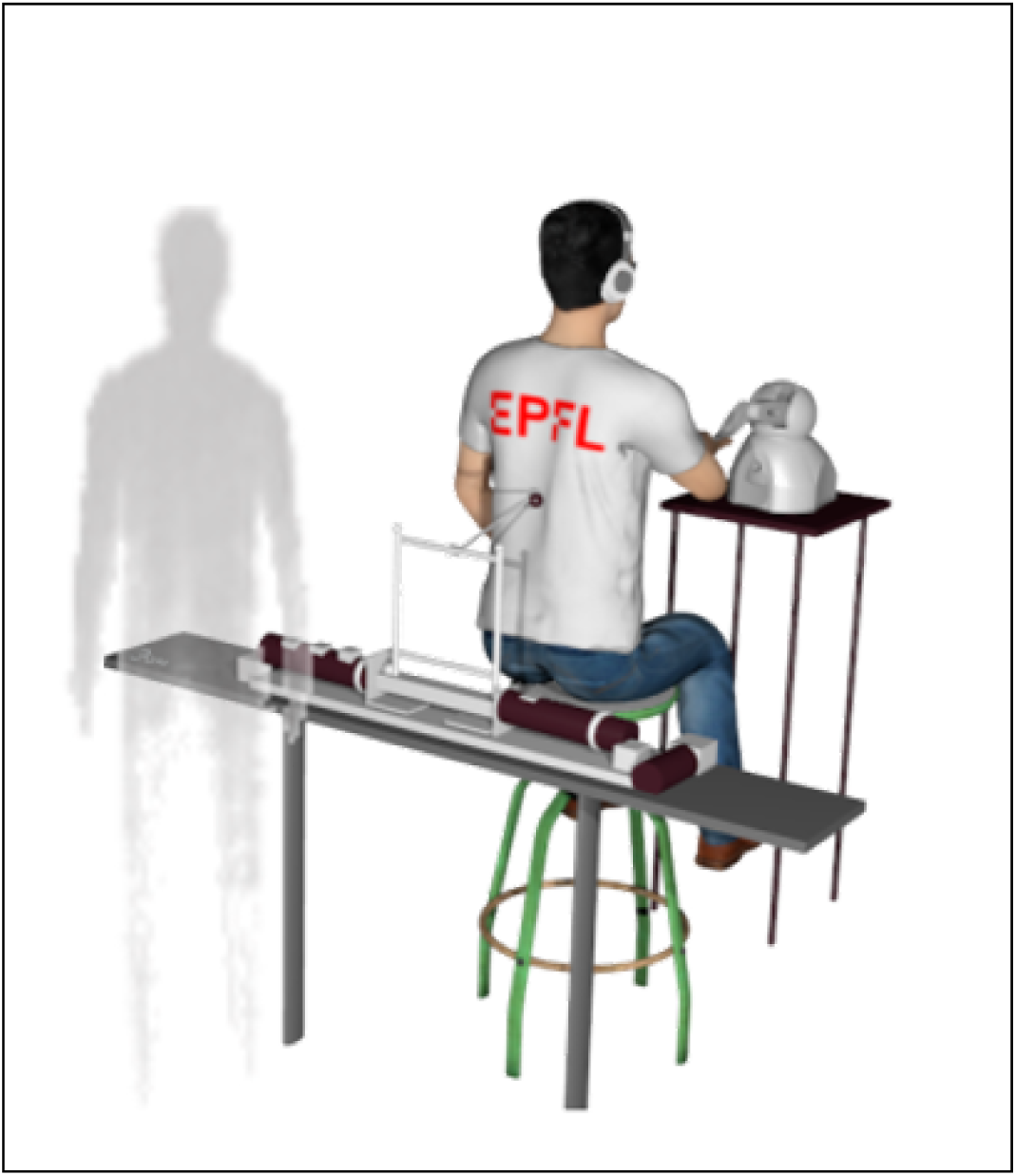
Robotic somatomotor stimulation task. Participant using the robotic system with a front robot and a back robot. A robot-induced PH (riPH) is schematically shown behind the participant.

### Statistical analysis

All statistical analyses were conducted using R (version 4.5.0) within RStudio (version 2025.05.0). To compare the PD-MH and PD-nH groups in terms of socio-demographic characteristics, clinical variables, and scores from the neuropsychological tests, independent Welch’s t-test were performed. For the robotic somatomotor stimulation task, data from 29 PD-MH and 37 PD-nH patients was analyzed, as 2 patients from the PD-MH group did not complete the task due to severe dyskinesia. A generalized linear mixed-effects model (glmm) with a binomial link function (glmer) was used to analyze patients’ sensitivity to riPH at the trial level, with each individual trial response (“Yes vs. “No” to index the global riPH sensitivity for each subject) as the dependent variable. The model included group (two factors: PD-MH vs PD-nH) and standardized delay of asynchrony (0, 250, 500 ms) as fixed effects, as well as their interaction. A random intercept for each participant was included to account for the non-independence of repeated measures. Contrast sum was used for the categorical variables in all the analyses performed. Delay was standardized (z-scored) to improve model convergence and interpretability by placing all predictors on a comparable scale. To examine the relationship between neuropsychological performance and sensitivity to riPH, we conducted similar series of GLMMs with a binomial link function. In each model for each neuropsychological score, the dependent variable was the binary responses to riPH, with random intercepts for participants to account for repeated measures across delay conditions (0, 250, 500 ms). Group (PD-MH vs. PD-nH) was included as a categorical factor and z-scores from each neuropsychological test served as continuous predictors. These included the frontal-subcortical and posterior cortical from the PD-CRS; the number of self-corrected and non-corrected errors on the TMT parts A and B; the number of self-corrected and non-corrected errors on the Stroop test (conditions 3 and 4) and the SCI scores. Contrast sum coding was applied for the categorical variables in all the analyses performed. Z-scoring was applied to the cognitive scores to standardize them (mean=0, standard deviation=1), ensuring they were on a comparable scale and improving GLMM convergence and interpretability particularly when combining predictors with different units of measurement. By standardizing the scores, we also accounted for potential differences in the variability and range of each test, facilitating a clearer comparison of their contributions to the model. Interaction terms between group and each cognitive variable were included to test whether the relationship between cognitive performance and riPH sensitivity differed by group. When significant interaction effects were observed, post-hoc group-specific GLMMs were conducted to examine how cognitive performance or scores and delay influenced riPH sensitivity within each group.

To ensure that the observed relationships between riPH global sensitivity and cognitive performance were not confounded by clinical or demographic variables, additional generalized linear mixed-effects models were run including covariates of no interest: age, sex, education, disease duration, LEDD, MDS-UPDRS III scores, and depression and anxiety scores. These adjusted models tested whether the interaction between group (PD-MH vs. PD-nH), delay and cognitive variables remained significant after accounting for these covariates, confirming that the associations were not explained by other clinical or demographic factors. Statistical significance was set at *P*<0.05.

Bonferroni correction was applied for ten planned comparisons involving the following predictors: frontal subcortical and posterior cortical scores of the PD-CRS scores; self-corrected and non-corrected errors of Stroop test (conditions 3 and 4), non-corrected errors of TMT A, self-corrected and non-corrected errors of TMT parts B and SCI, each interacting with subgroup and delay.

## Results

### Clinical evaluation

In the PD-MH group, passage hallucinations were the most common (61.29%), followed by visual illusions/misperceptions (58.06%) and presence hallucinations (54.84%). The different types of MH and their co-occurrence among the PD-MH group are illustrated in the Supplemental Material 2 (**Figure S1)**.

There was no significant difference between the PD-MH and PD-nH groups in terms of age, sex, education, illness duration and LEDD (all *P*s>0.05). On the MDS-UPDRS scale, only non-motor symptoms (MDS-UPDRS I; t=3.54; df=59.25; *P*<0.001) were rated as significantly more severe by the PD-MH group than the PD-nH group; the other subscales assessing activities of daily life (MDS-UPDRS II), severity of motor symptoms (MDS-UPDRS III) and motor complications (MDS-UPDRS IV) as well as Hoehn and Yahr staging of severity of PD were not significantly different between groups (all *P*s>0.05). Additionally, PD-MH patients scored significantly higher on depression (t=2.12; df=54.71; *P*=0.04) and anxiety (t=2.29; df=60.69; *P*=0.03) compared to the PD-nH group, as measured by the HADS. However, the mean scores of depression and anxiety in both groups were below the pathological cut-off (score <7), indicating the absence of clinically relevant symptomatology in both groups (**Table 1**).

### Neuropsychological evaluation

Analyses of behavioral performance on neuropsychological tests assessing general cognition (MoCA), frontal subcortical and posterior cortical functions (PD-CRS), as well as cognitive flexibility, resistance to interference and inhibitory control (number of errors in TMT-A and B and in the Stroop color-word test) did not reveal any significant group differences between PD-MH and PD-nH patients (all *P*s>0.05). The total scores on the MoCA and PD-CRS indicated preserved cognitive functioning, falling above the established thresholds for both dementia and mild cognitive impairment (see **Table 2**). However, PD-MH patients reported significantly higher subjective cognitive decline or complaints in daily life compared to PD-nH patients (t=3.87; df=50.77; *P*<0.001). A detailed summary of the neuropsychological results for both groups is presented in **Table 2**.

**Table 2.**
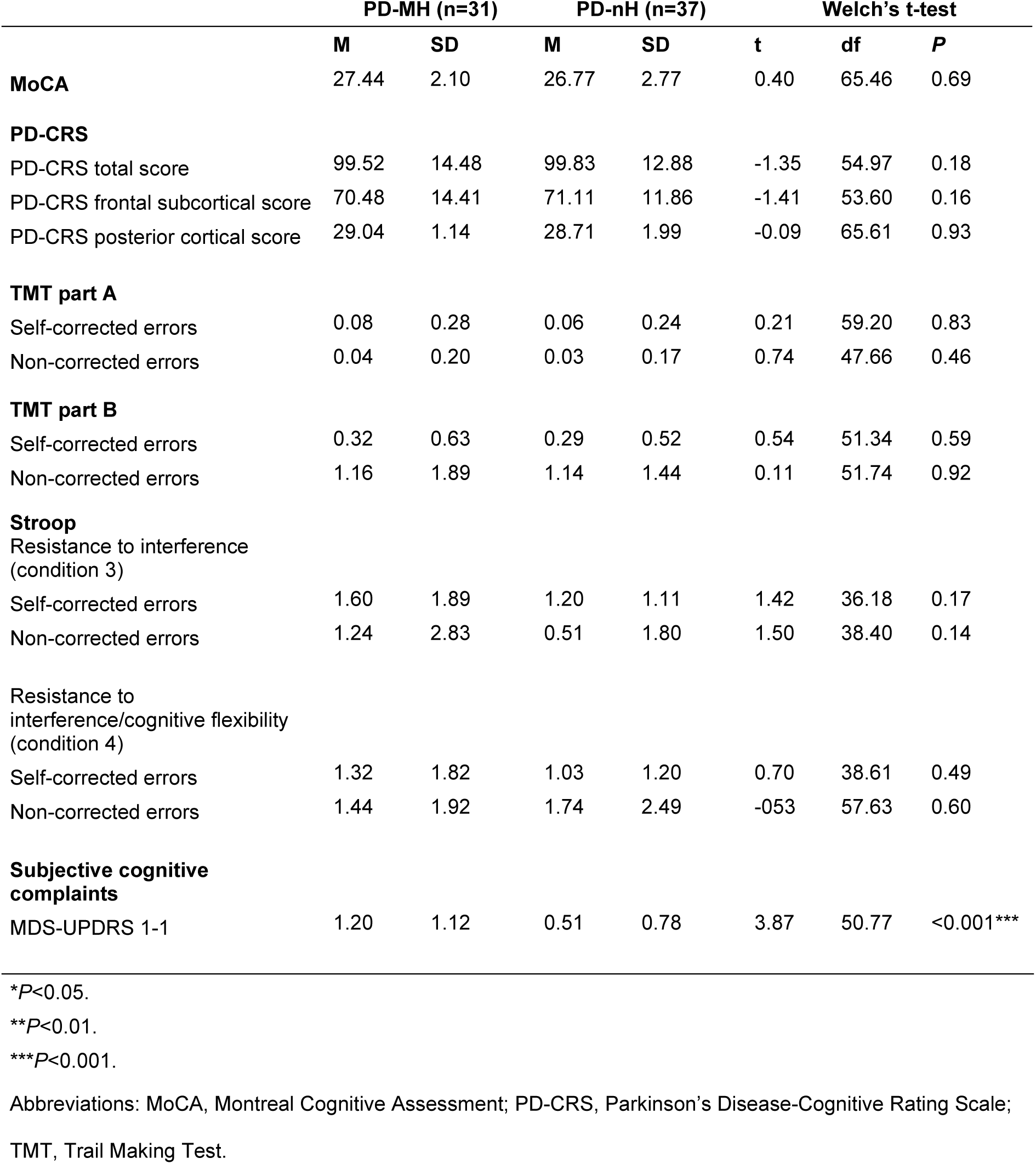
Neuropsychological score differences between PD patient groups.

|  | PD-MH (n=31) |  | PD-nH (n=37) |  | Welch's t-test |  |  |
| --- | --- | --- | --- | --- | --- | --- | --- |
|  | M | SD | M | SD | t | df | P |
| **MoCA** | 27.44 | 2.10 | 26.77 | 2.77 | 0.40 | 65.46 | 0.69 |
| **PD-CRS** |  |  |  |  |  |  |  |
| PD-CRS total score | 99.52 | 14.48 | 99.83 | 12.88 | -1.35 | 54.97 | 0.18 |
| PD-CRS frontal subcortical score | 70.48 | 14.41 | 71.11 | 11.86 | -1.41 | 53.60 | 0.16 |
| PD-CRS posterior cortical score | 29.04 | 1.14 | 28.71 | 1.99 | -0.09 | 65.61 | 0.93 |
| **TMT part A** |  |  |  |  |  |  |  |
| Self-corrected errors | 0.08 | 0.28 | 0.06 | 0.24 | 0.21 | 59.20 | 0.83 |
| Non-corrected errors | 0.04 | 0.20 | 0.03 | 0.17 | 0.74 | 47.66 | 0.46 |
| **TMT part B** |  |  |  |  |  |  |  |
| Self-corrected errors | 0.32 | 0.63 | 0.29 | 0.52 | 0.54 | 51.34 | 0.59 |
| Non-corrected errors | 1.16 | 1.89 | 1.14 | 1.44 | 0.11 | 51.74 | 0.92 |
| **Stroop** |  |  |  |  |  |  |  |
| Resistance to interference (condition 3) |  |  |  |  |  |  |  |
| Self-corrected errors | 1.60 | 1.89 | 1.20 | 1.11 | 1.42 | 36.18 | 0.17 |
| Non-corrected errors | 1.24 | 2.83 | 0.51 | 1.80 | 1.50 | 38.40 | 0.14 |
| Resistance to interference/cognitive flexibility (condition 4) |  |  |  |  |  |  |  |
| Self-corrected errors | 1.32 | 1.82 | 1.03 | 1.20 | 0.70 | 38.61 | 0.49 |
| Non-corrected errors | 1.44 | 1.92 | 1.74 | 2.49 | -0.53 | 57.63 | 0.60 |
| **Subjective cognitive complaints** |  |  |  |  |  |  |  |
| MDS-UPDRS 1-1 | 1.20 | 1.12 | 0.51 | 0.78 | 3.87 | 50.77 | <0.001*** |

\* $P<0.05$ .
\*\* $P<0.01$ .
\*\*\* $P<0.001$ .
Abbreviations: MoCA, Montreal Cognitive Assessment; PD-CRS, Parkinson's Disease-Cognitive Rating Scale; TMT, Trail Making Test.

### Robot-induced presence hallucinations (riPH)

We found that the PD-MH group had a significantly higher riPH sensitivity (indexed by a higher proportion of affirmative responses in the PH-inducing robotic procedure) than the PD-nH group, as indicated by a main effect of group in the generalized linear mixed model (β=1.81, SE=0.70, z=2.58, *P*=0.01) (**Figure 2**). A main effect of delay was also observed (β=0.65, SE=0.09, z=6.96, *P*<0.001), indicating that longer delays increased the likelihood of affirmative responses to riPH. Consistent with previous findings [36], this result indicates that PD-MH patients showed higher sensitivity to conflicting somatomotor stimulation than PD-nH patients. No significant interaction was observed between group and delay (β=0.01, SE=0.09, z=0.13, P=0.90), suggesting that the effect of delay on riPH sensitivity did not vary by group. Random intercepts were included to account for repeated measures across participants.

**Figure 2.**
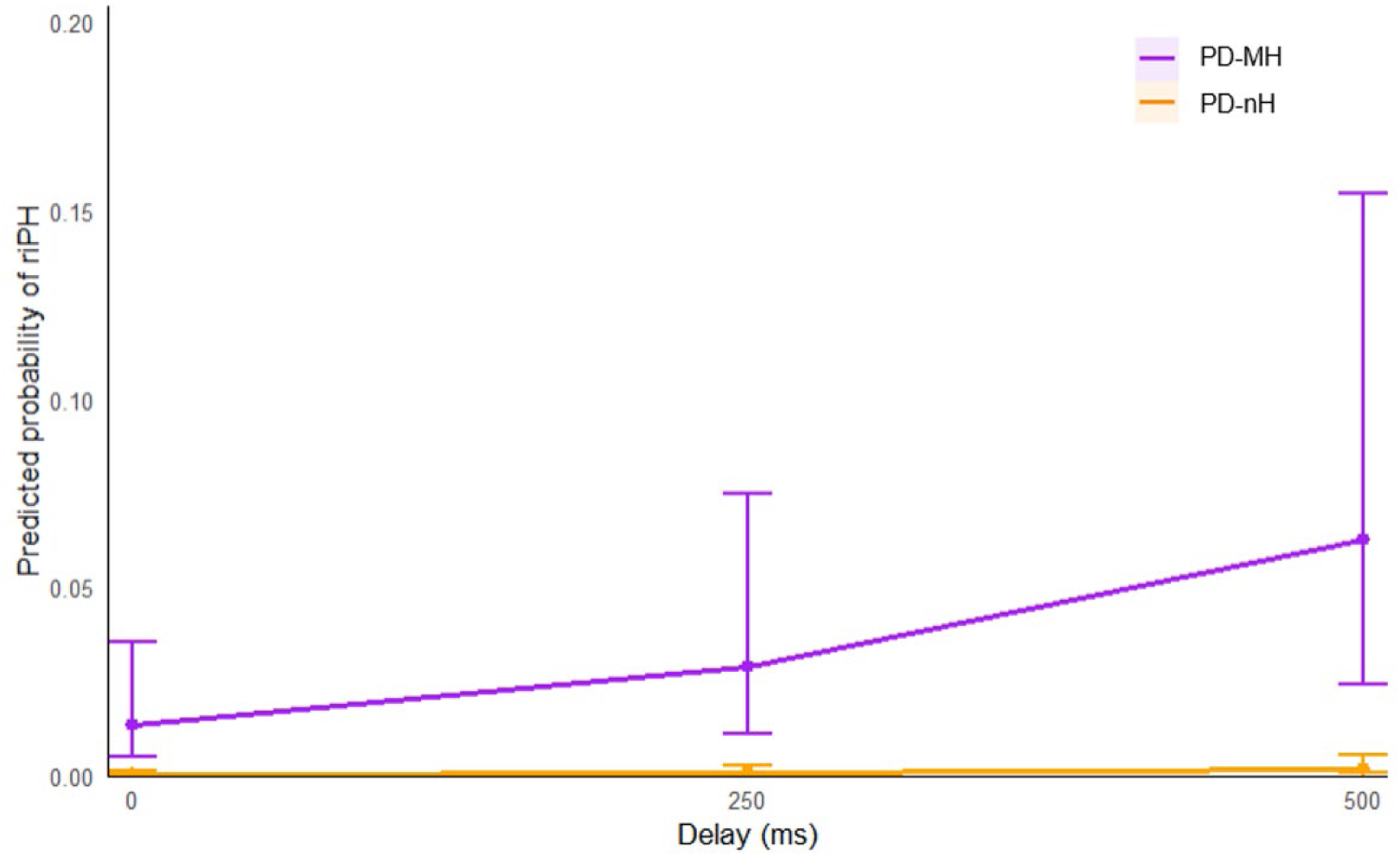
Predicted probability of robot-induced presence hallucination (riPH) as a function of delay in somatomotor stimulation. Predicted probabilities were estimated from a GLMM including z-scored delays and subgroup as fixed effects, and random intercepts for participants. In PD-MH patients (purple line), the probability of riPH increases with longer delays, whereas in PD-nH patients (orange line), the predicted probability remains relatively low across delays. Points indicate predicted probabilities at each discrete delay, and error bars represent ±1 standard error (SE) around the model-based predictions.

### Relationship between riPH and neuropsychology PD-CRS

We first examined whether cognitive performance modulated sensitivity to riPH as a function of group and delay. The results revealed a significant three-way interaction between group, frontal subcortical scores, and delay (β=0.29, SE=0.10, z=2.74, *P*=0.006), indicating that the relationship between frontal subcortical cognitive performance and riPH sensitivity differed across groups and levels of delay asynchrony.

Post hoc, group-specific analyses showed that in the PD-MH group, lower frontal subcortical scores (i.e., greater cognitive alteration) were associated with a higher probability of riPH (β=−1.50, SE=0.69, z=−2.18, *P*=0.03), together with a significant main effect of delay (β=0.69, SE=0.13, z=5.39, *P*<0.001) (**Figure 3**). The interaction between frontal subcortical scores and delay was not significant (*P*=0.25), indicating that frontal subcortical deficits increased riPH sensitivity similarly across delay conditions.

**Figure 3.**
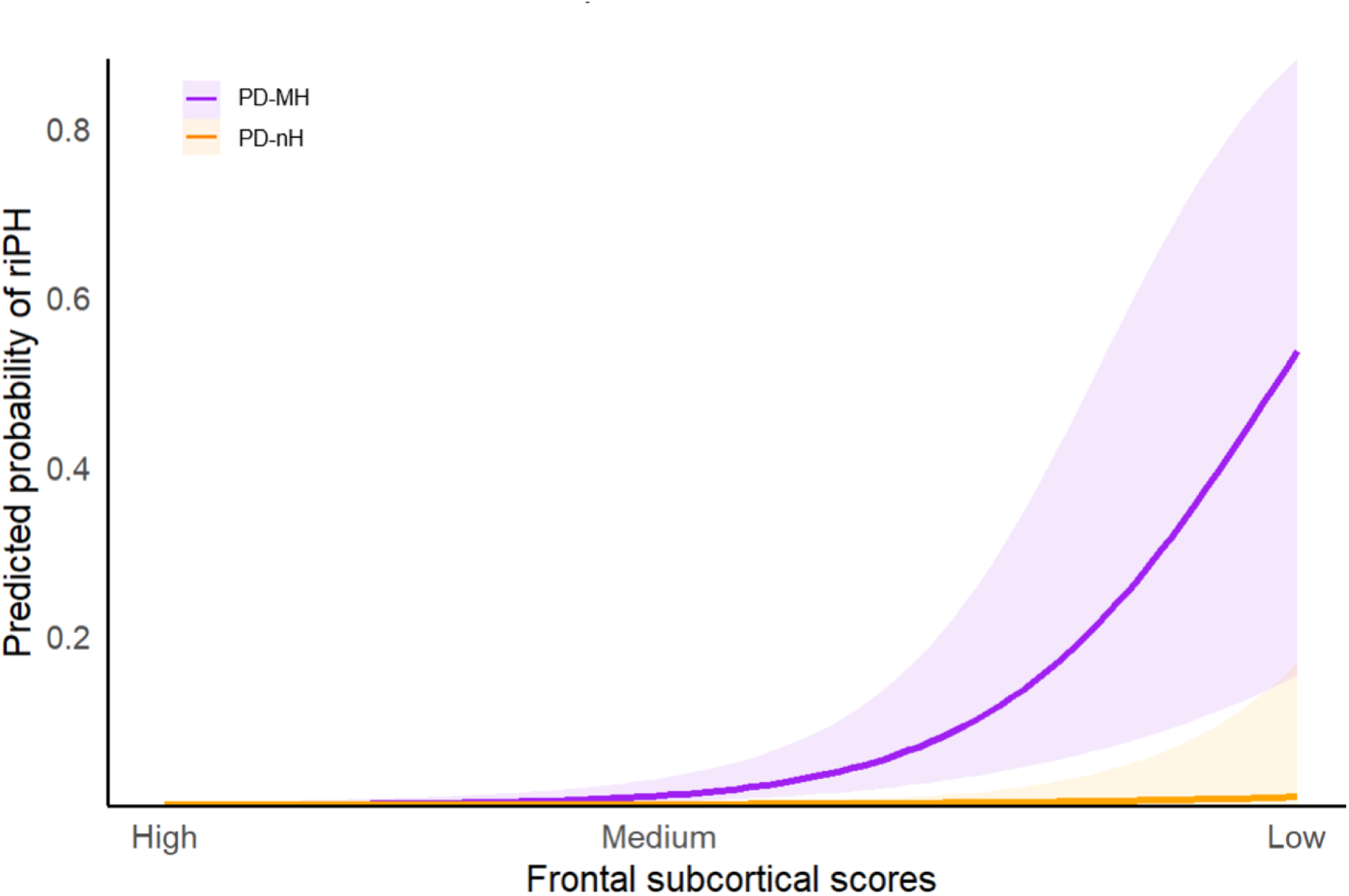
Relationship between PD-CRS frontal subcortical scores and predicted probability of robot-induced presence hallucination (riPH) by group. Lower frontal subcortical scores (indicating impaired cognitive performance) are associated with higher riPH probability. This effect was significant in PD-MH patients (purple line, *P*=0.03), whereas in PD-nH patients (orange line), predicted probability remains largely constant across PD-CRS scores (*P*=0.71). Shaded areas represent ±1 standard error (SE) around model-based predictions.

**Figure 4.**
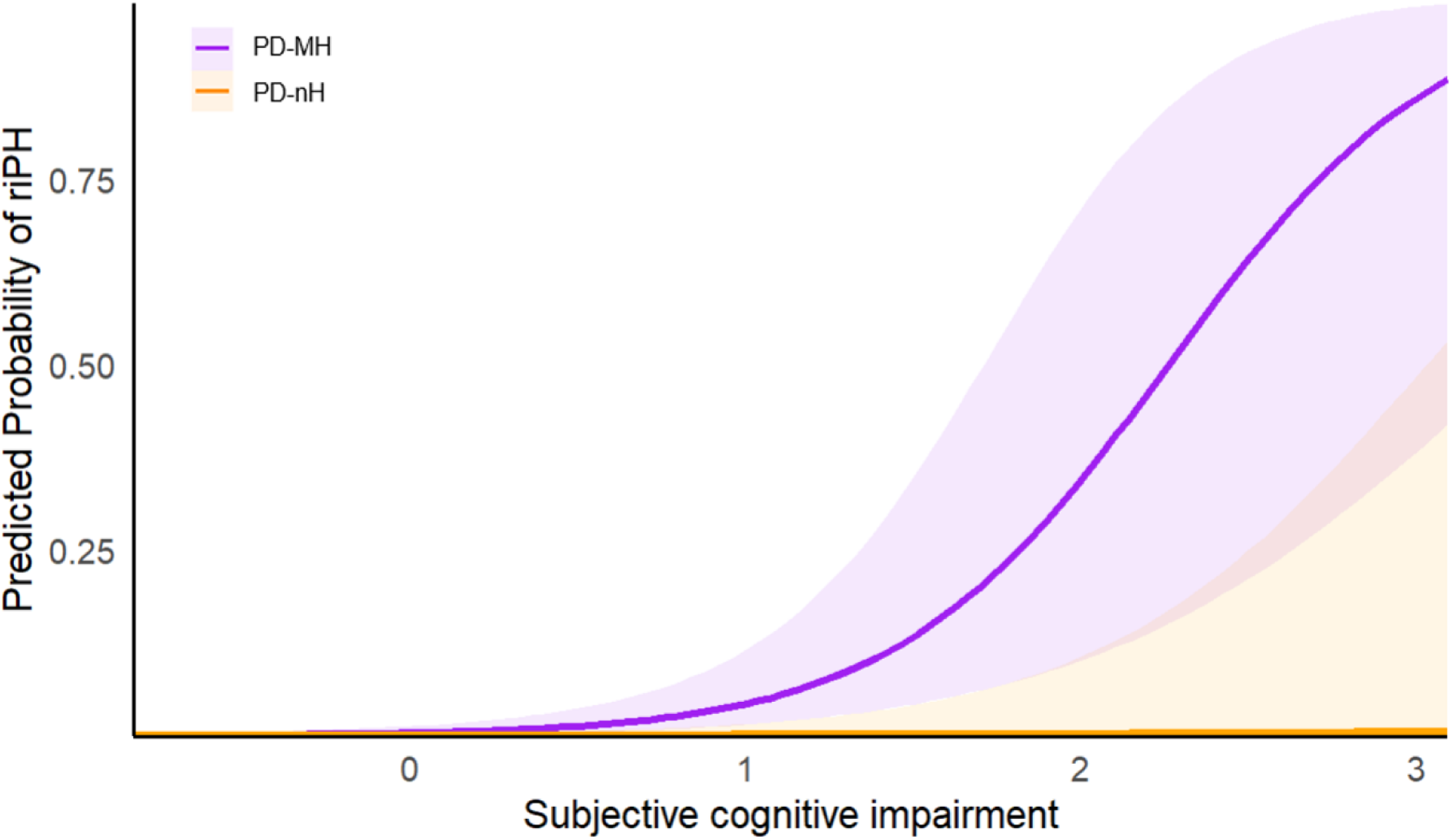
Predicted probability of robot-induced presence hallucination (riPH) by subjective cognitive impairment (SCI) in PD-MH and PD-nH patients. Higher SCI ratings (indicating worse cognitive complaints) are associated with a significantly increased probability of riPH in PD-MH patients (purple line, *P*=0.008), whereas SCI is not significantly associated with riPH in PD-nH patients (orange line, *P*=0.77), and variability around predictions is minimal. Shaded areas indicate ±1 standard error (SE) around model-based predictions.

In contrast, in the PD-nH group, frontal subcortical scores showed no main effect on riPH sensitivity (β=−0.46, SE=1.24, z=−0.37, *P*=0.71), while riPH sensitivity increased with delay (β=0.61, SE=0.14, z=4.23, *P*<0.001). A significant interaction between frontal subcortical scores and delay (β=−0.48, SE=0.19, z=−2.53, *P*=0.01), indicated that frontal subcortical performance modestly influenced riPH sensitivity at longer delays.

Together, these results indicate that in PD-MH patients, frontal subcortical cognitive alteration constitutes a general susceptibility factor for riPH across all delay conditions, whereas in PD-nH patients, riPH sensitivity is largely influenced by delay.

### Subjective cognitive impairment (SCI)

We also examined whether SCI (MDS-UPDRS I, item 1.1) modulated patients’ sensitivity to riPH as a function of group and delay. The results showed a significant three-way interaction between group, ratings of SCI, and delay (β=−0.35, SE=0.15, z=−2.38, *P*=0.017), indicating that the relationship between SCI and riPH sensitivity differed across groups and levels of delay. Post hoc, group-specific models showed that in the PD-MH group, higher levels of SCI were associated with a higher sensitivity to riPH (main effect of SCI: β=1.99, SE=0.76, z=2.64, *P*=0.008), and riPH sensitivity also increased with delay (main effect of delay: β=1.27, SE=0.21, z=6.14, *P*<0.001), although the influence of SCI diminished at longer delays (interaction between SCI and delay: β=−0.555, SE=0.127, z=−4.35, *P*<0.001).

In the PD-nH group, riPH sensitivity increased with delay (β=0.68, SE=0.15, z=4.45, *P*<0.001), while SCI had no significant effect (main or interaction; all *P*>0.7). These results indicate that SCI modulates riPH sensitivity in PD-MH patients, but not in PD-nH patients.

The relationship between these measures (frontal subcortical scores; SCI) with delay and group on the sensitivity to riPH remained significant after controlling for key clinical and demographic variables (covariate of non-interest) such as age, education, sex, disease duration, LEDD, motor symptoms severity, and mood disturbances (depression and anxiety) (all *Ps*<0.05).

## Discussion

The main objective of this study was to determine whether MH, one of the most frequent and early-occurring hallucinations in PD, are associated with cognitive deficits and how this potential relationship depends on sensitivity to the riPH procedure. For this, we combined in depth neurological and neuropsychological examinations with an experimental approach during which PH are induced in patients by means of a robotic device delivering conflicting somatomotor stimulations [36, 39, 40]. This extends earlier studies, which analyzed the relationship between hallucinations and neuropsychology by classifying PD patients only based on interviews or self-reports about having or not hallucinations. By providing an additional real-time measure of PH susceptibility, this approach may allow for the detection of more subtle cognitive vulnerabilities linked to MH. Consistent with our hypothesis, we report (1) a significant association between increased sensitivity to riPH and frontal subcortical deficits. Critically, this relationship was particularly (2) strong in the PD-MH group where lower frontal subcortical scores (i.e., greater cognitive deficits) were associated with higher riPH sensitivity and differed from PD-nH patients. In contrast, (3) no significant association was found between riPH sensitivity and posterior cortical functions. Moreover, (4) we found a significant relationship between increased sensitivity to riPH and higher SCI reported by patients, especially in the MH group; and (5) these associations were independent of clinical and demographic differences between groups. These findings provide evidence that MH are associated with ‘minor’ frontal subcortical executive vulnerability in PD [37, 44], and we argue that the present approach - the combination of the riPH procedure with neurological and neuropsychological examinations - allows the quantification of this more subtle relationship between MH, riPH, and frontal (executive) dysfunction that standard clinical assessments may fail to capture.

While both frontal subcortical and posterior cortical dysfunctions have been consistently associated with structured visual hallucinations [13, 25, 56], the cognitive correlates of MH remained elusive. This is likely due to the lack of sensitive assessment methods capable of detecting more subtle frontal subcortical executive deficits in PD patients with MH. Accordingly, PD-MH versus PD-nH groups often show only small and non-significant differences on standard neuropsychological tests [8, 38, 57]. In the present study, we also found comparable neuropsychological performance between PD-MH and PD-nH groups on frontal subtests [7, 8, 35]. However, when considering sensitivity to riPH, two distinct patterns emerged that clarify the role of frontal subcortical executive functions across groups. First, lower frontal subcortical scores were associated with higher riPH sensitivity, indicating that greater frontal subcortical executive alteration relates to increased susceptibility to robotic somatomotor stimulation. Second, this association differed between groups, being more robust and delay-independent in PD-MH patients, whereas in PD-nH patients, frontal subcortical performance primarily modulated how riPH sensitivity increased with delay. These findings extend prior research implicating frontal lobe dysfunction in MH [7, 34, 35, 58] and demonstrate that additional measures such as sensitivity to riPH are important to detect these subtle deficits. The riPH measure is based on exposure to conflicting somatomotor signals (tactile, proprioceptive, motor) and has been shown to recruit a frontal subcortical network consisting of primary somatomotor cortex, premotor cortex, supplementary motor cortex and basal ganglia [36]. Combining riPH sensitivity with comprehensive standard executive functions tests associated with frontal subcortical circuits as included in the PD-CRS, we distinguished PD-MH from PD-nH by revealing a neuropsychological deficit that was undetected by the conventional neuropsychological assessments that we carried out.

Importantly, frontal subcortical executive deficits captured by the PD-CRS subscale provide a composite measure integrating multiple domains (e.g., attention, working memory, immediate and delayed recall, verbal fluencies) [38, 59, 60] and appear more representative of subtle or minor cognitive deficits in PD-MH patients than single-dimension neuropsychological tests. Such multi-dimensional assessments of executive functions, especially when combined with riPH sensitivity measure, can uncover early cognitive vulnerabilities that are more likely to be missed by single tests focusing on one aspect of executive functions. Notably, these associations were independent of clinical variables such as motor severity, disease duration, medication level, or mood, as well as sociodemographic factors including age, sex, and education.

Taken together, these findings show that the combination of riPH sensitivity with neuropsychology allows to describe a subtle ‘minor’ frontal subcortical executive deficit in PD-MH patients, independent of posterior brain functions. Thus, riPH sensitivity was not associated with cognitive domains related to posterior cortical functioning, neither in the PD-MH nor the PD-nH group. We argue accordingly that posterior functions and related brain regions, which are typically altered in PD patients with structured visual hallucinations and associated with later cognitive decline or dementia [19, 27, 30], are still preserved at the MH stage of PD. These findings support the idea that, although MH have been consistently associated with structural and functional deficits in posterior visuo-perceptive brain regions [7, 61], the clinical manifestation of MH depends also and importantly on the degree of frontal subcortical dysfunction [7, 30, 44]. Overall, the present results support the idea that MH may serve as an early indicator of cognitive vulnerability in PD, particularly when assessed alongside riPH measures.

Additionally, PD-MH patients reported significantly greater SCI in daily life (MDS-UPDRS I, item 1.1) compared to PD-nH patients and that riPH sensitivity was significantly modulated by SCI in PD-MH but not in PD-nH patients. This supports the idea that SCI may reflect a cognitive mental state associated with increased sensitivity to conflicting somatomotor stimulation. Although SCI is not the primary indicator of cognitive vulnerability, it provides complementary information to objective measures such as the PD-CRS frontal subcortical scale and riPH sensitivity. Therefore, SCI – when considered alongside neuropsychological testing and riPH sensitivity measurements – should receive greater attention in clinical settings [24, 35, 62], as it may indicate early cognitive changes that remain undetected by standard neuropsychological testing in patients with normal or near-normal performance. Overall, these findings suggest that patients with MH could benefit from careful follow-up and, when available, testing with riPH measures to better identify and manage emerging frontal executive cognitive impairments [63–65]. Thus, MH could serve as an early indicator of cognitive alteration in PD [38, 45, 66], particularly when combined with riPH measurements and complementary assessments such as SCI.

These relationships between riPH sensitivity and cognitive measures cannot be explained by differences in clinical or demographic variables. Although PD-MH patients had higher anxiety and depression levels (still below clinical threshold) and greater severity of non-motor symptoms of daily living (MDS-UPDRS I), the main findings remained independent of motor symptom severity (MDS-UPDRS III), disease duration, medication level (LEDD), and mood disturbances, which have previously been associated with hallucinations and/or cognitive deficits [1, 2, 57, 67–70]. Furthermore, the relationship between frontal subcortical alterations and riPH sensitivity was not attributable to sociodemographic factors such as age, sex or level of education.

The present study has several limitations. First, our study being cross-sectional, it does not allow us to capture the progression of the frontal subcortical executive (and other) impairments of PD-MH patients over time. Second, since PD-nH patients did not experience hallucinations at the time of the study, some may develop them later, as observed in previous longitudinal studies [71–73]. In line with this, we found that several PD-nH patients already showed higher riPH sensitivity associated with poorer frontal subcortical performance. These PD-nH patients may represent an early vulnerability [35, 38] and could be at higher risk of developing MH (and later structured visual hallucinations), highlighting the need for closer follow-up in future studies. Finally, we assessed SCI with a single question from the MDS-UPDRS scale, but upcoming studies would benefit from more comprehensive questionnaires [74, 75] to evaluate subjective cognitive decline.

In conclusion, our study demonstrates that PD-MH, compared to PD-nH, is associated with stronger frontal subcortical executive deficits that depend on riPH sensitivity. Future brain imaging and electrophysiological studies should investigate the neural mechanisms underlying this association and longitudinal research will be important to describe the clinical and neuropsychological trajectory of PD-MH patients over time.

## Acknowledgement

The authors would like to express their sincere gratitude to all the patients for their valuable time, effort, and for sharing their personal experiences with hallucinations. We also extend our thanks to Dr. Atena Fadaei Jouybari for her contribution to the design of **Figure 1**.

Study data were collected and managed using REDCap electronic data capture tools hosted at the Swiss Federal School of Technology (Ecole Polytechnique Fédérale de Lausanne). REDCap (Research Electronic Data Capture) is a secure, web-based software platform designed to support data capture for research studies, providing 1) an intuitive interface for validated data capture; 2) audit trails for tracking data manipulation and export procedures; 3) automated export procedures for seamless data downloads to common statistical packages; and 4) procedures for data integration and interoperability with external sources.

## Author Contributions (CRediT)

**J.P.:** Conceptualization; Methodology; Validation; Formal analysis; Investigation; Resources; Data curation; Writing - original draft; Writing - review & editing; Visualization; Supervision; Project administration; Funding acquisition. **L.F.D.P.T:** Validation; Investigation; Writing - review & editing; Project administration. **F.B.:** Methodology; Formal analysis; Writing – review & editing; Funding acquisition. **N.H.M.:** Formal analysis; Writing – review & editing. **L.J.:** Software; writing – review & editing. **M.E.M-G.:** Investigation; Writing - review & editing; Project administration. **C.S.**: Investigation; Writing - review & editing. **S.C.C:** Investigation; Writing - review & editing. **J.F.B.:** Resources; Writing - review & editing. **M.C.J.:** Resources; Writing - review & editing. **V.F.:** Resources; Writing - review & editing. **J.H.:** Resources; Writing - review & editing. **B.W.:** Resources; Writing - review & editing. **J.P.M.:** Methodology; Writing - review & editing. **P.K.:** Methodology; Writing - review & editing. **O.B.:** Conceptualization; Methodology; Resources; Writing - review & editing; Visualization; Supervision; Funding acquisition.

## Statements and declarations

### Ethical considerations

The study was approved by the cantonal ethics committees of Geneva, Vaud and Bern (Switzerland) under protocol numbers 2019-02275 and 2021-01608.

### Consent to participate

All participants were presented to the study orally and by means of a written information sheet. All participants gave their written informed consent prior enrollment in the study.

### Consent for publication

Not applicable

## Data availability

All data produced in the present study are available upon reasonable request to the authors.

## References

[1] Fénelon G, Mahieux F, Huon R, et al. Hallucinations in Parkinson’s disease: prevalence, phenomenology and risk factors. Brain 2000; 123 ( Pt 4): 733–745.

[2] Ffytche DH, Pereira JB, Ballard C, et al. Risk factors for early psychosis in PD: insights from the Parkinson’s Progression Markers Initiative. J Neurol Neurosurg Psychiatr 2017; 88: 325–331.

[3] Mack J, Rabins P, Anderson K, et al. Prevalence of psychotic symptoms in a community-based Parkinson disease sample. Am J Geriatr Psychiatry 2012; 20: 123–132.

[4] Ffytche DH, Creese B, Politis M, et al. The psychosis spectrum in Parkinson disease. Nat Rev Neurol 2017; 13: 81–95.

[5] Weintraub D, Aarsland D, Chaudhuri KR, et al. The neuropsychiatry of Parkinson’s disease: advances and challenges. The Lancet Neurology 2022; 21: 89–102.

[6] Albert L, Vehar N, Potheegadoo J, et al. Visual hallucinations in Parkinson’s disease are associated with deficits in social perception. Journal of Parkinson’s Disease 2025; 1877718X251336196.

[7] Pagonabarraga J, Bejr-Kasem H, Martinez-Horta S, et al. Parkinson disease psychosis: from phenomenology to neurobiological mechanisms. Nat Rev Neurol 2024; 1–16.

[8] Llebaria G, Pagonabarraga J, Martínez-Corral M, et al. Neuropsychological correlates of mild to severe hallucinations in Parkinson’s disease. Mov Disord 2010; 25: 2785–2791.

[9] Diederich NJ, Fénelon G, Stebbins G, et al. Hallucinations in Parkinson disease. Nat Rev Neurol 2009; 5: 331–342.

[10] Aarsland D, Larsen JP, Tandberg E, et al. Predictors of nursing home placement in Parkinson’s disease: a population-based, prospective study. J Am Geriatr Soc 2000; 48: 938–942.

[11] Goetz CG, Stebbins GT. Risk factors for nursing home placement in advanced Parkinson’s disease. Neurology 1993; 43: 2227–2229.

[12] Fénelon G, Soulas T, Zenasni F, et al. The changing face of Parkinson’s disease-associated psychosis: a cross-sectional study based on the new NINDS-NIMH criteria. Mov Disord 2010; 25: 755–759.

[13] Santangelo G, Trojano L, Vitale C, et al. A neuropsychological longitudinal study in Parkinson’s patients with and without hallucinations. Movement Disorders 2007; 22: 2418–2425.

[14] Pisani S, Gosse L, Aarsland D, et al. Parkinson’s disease psychosis associated with accelerated multidomain cognitive decline. BMJ Ment Health 2024; 27: 1–10.

[15] Factor SA, Scullin MK, Sollinger AB, et al. Cognitive correlates of hallucinations and delusions in Parkinson’s disease. J Neurol Sci 2014; 347: 316–321.

[16] Emre M, Aarsland D, Brown R, et al. Clinical diagnostic criteria for dementia associated with Parkinson’s disease. Mov Disord 2007; 22: 1689–1707; quiz 1837.

[17] Litvan I, Goldman JG, Tröster AI, et al. Diagnostic criteria for mild cognitive impairment in Parkinson’s disease: Movement Disorder Society Task Force guidelines. Mov Disord 2012; 27: 349–356.

[18] Poewe W, Gauthier S, Aarsland D, et al. Diagnosis and management of Parkinson’s disease dementia. Int J Clin Pract 2008; 62: 1581–1587.

[19] Devignes Q, Viard R, Betrouni N, et al. Posterior Cortical Cognitive Deficits Are Associated With Structural Brain Alterations in Mild Cognitive Impairment in Parkinson’s Disease. Frontiers in Aging Neuroscience; 13, https://www.frontiersin.org/article/10.3389/fnagi.2021.668559 (2021, accessed 20 February 2022).

[20] Chandler JM, Nair R, Biglan K, et al. Characteristics of Parkinson’s Disease in Patients with and without Cognitive Impairment. Journal of Parkinson’s Disease 2021; 11: 1381–1392.

[21] Weil RS, Costantini AA, Schrag AE. Mild Cognitive Impairment in Parkinson’s Disease—What Is It? Curr Neurol Neurosci Rep 2018; 18: 17.

[22] Biundo R, Bezdicek O, Cammisuli DM, et al. Attention/Working Memory and Executive Function in Parkinson’s Disease: Review, Critique, and Recommendations. Movement Disorders; n/a. Epub ahead of print 2025. DOI: 10.1002/mds.30293.

[23] Williams DR, Warren JD, Lees AJ. Using the presence of visual hallucinations to differentiate Parkinson’s disease from atypical parkinsonism. J Neurol Neurosurg Psychiatry 2008; 79: 652–655.

[24] Santos-García D, de Deus Fonticoba T, Cores Bartolomé C, et al. Risk of Cognitive Impairment in Patients With Parkinson’s Disease With Visual Hallucinations and Subjective Cognitive Complaints. J Clin Neurol.

[25] Pezzoli S, Sánchez-Valle R, Solanes A, et al. Neuroanatomical and cognitive correlates of visual hallucinations in Parkinson’s disease and dementia with Lewy bodies: Voxel-based morphometry and neuropsychological meta-analysis. Neurosci Biobehav Rev 2021; 128: 367–382.

[26] Kehagia AA, Barker RA, Robbins TW. Neuropsychological and clinical heterogeneity of cognitive impairment and dementia in patients with Parkinson’s disease. Lancet Neurol 2010; 9: 1200–1213.

[27] Puig-Davi A, Martinez-Horta S, Pérez-Carasol L, et al. Prediction of Cognitive Heterogeneity in Parkinson’s Disease: A 4-Year Longitudinal Study Using Clinical, Neuroimaging, Biological and Electrophysiological Biomarkers. Annals of Neurology; n/a. Epub ahead of print 5 August 2024. DOI: 10.1002/ana.27035.

[28] Barnes J, Boubert L. Executive functions are impaired in patients with Parkinson’s disease with visual hallucinations. J Neurol Neurosurg Psychiatry 2008; 79: 190–192.

[29] Creese B, Albertyn CP, Dworkin S, et al. Executive function but not episodic memory decline associated with visual hallucinations in Parkinson’s disease. J Neuropsychol. Epub ahead of print 23 July 2018. DOI: 10.1111/jnp.12169.

[30] Pagonabarraga J, Kulisevsky J, Llebaria G, et al. Parkinson’s disease-cognitive rating scale: a new cognitive scale specific for Parkinson’s disease. Mov Disord 2008; 23: 998–1005.

[31] Ozer F, Meral H, Hanoglu L, et al. Cognitive impairment patterns in Parkinson’s disease with visual hallucinations. Journal of Clinical Neuroscience 2007; 14: 742–746.

[32] Grossi D, Trojano L, Pellecchia MT, et al. Frontal dysfunction contributes to the genesis of hallucinations in non-demented Parkinsonian patients. International Journal of Geriatric Psychiatry 2005; 20: 668–673.

[33] Pagonabarraga J, Martinez-Horta S, Fernández de Bobadilla R, et al. Minor hallucinations occur in drug-naive Parkinson’s disease patients, even from the premotor phase. Mov Disord 2016; 31: 45–52.

[34] Bejr-kasem H, Pagonabarraga J, Martínez-Horta S, et al. Disruption of the default mode network and its intrinsic functional connectivity underlies minor hallucinations in Parkinson’s disease. Movement Disorders 2019; 34: 78–86.

[35] Bejr-Kasem H, Sampedro F, Marín-Lahoz J, et al. Minor hallucinations reflect early gray matter loss and predict subjective cognitive decline in Parkinson’s disease. Eur J Neurol 2021; 28: 438–447.

[36] Bernasconi F, Blondiaux E, Potheegadoo J, et al. Robot-induced hallucinations in Parkinson’s disease depend on altered sensorimotor processing in fronto-temporal network. Sci Transl Med 2021; 13: eabc8362.

[37] Bejr-kasem H, Martínez-Horta S, Pagonabarraga J, et al. The role of attentional control over interference in minor hallucinations in Parkinson’s disease. Parkinsonism & Related Disorders 2022; 102: 101–107.

[38] Lenka A, Hegde S, Arumugham SS, et al. Cognitive Correlates of Visual and Minor Hallucinations in Parkinson’s Disease. Canadian Journal of Neurological Sciences 2023; 50: 44–48.

[39] Bernasconi F, Blondiaux E, Rognini G, et al. Neuroscience robotics for controlled induction and real-time assessment of hallucinations. Nat Protoc 2022; 1–24.

[40] Blanke O, Pozeg P, Hara M, et al. Neurological and robot-controlled induction of an apparition. Curr Biol 2014; 24: 2681–2686.

[41] Albert L, Potheegadoo J, Herbelin B, et al. Numerosity estimation of virtual humans as a digital-robotic marker for hallucinations in Parkinson’s disease. Nat Commun 2024; 15: 1905.

[42] Potheegadoo J, Dhanis H, Horvath J, et al. Presence Hallucinations during Locomotion in Patients with Parkinson’s Disease. Mov Disord Clin Pract 2022; 9: 127–129.

[43] Salomon R, Progin P, Griffa A, et al. Sensorimotor Induction of Auditory Misattribution in Early Psychosis. Schizophrenia Bulletin 2020; sbz136.

[44] Bernasconi F, Pagonabarraga J, Bejr-Kasem H, et al. Theta oscillations and minor hallucinations in Parkinson’s disease reveal decrease in frontal lobe functions and later cognitive decline. Nat Mental Health 2023; 1: 477–488.

[45] Lenka A, George L, Arumugham SS, et al. Predictors of onset of psychosis in patients with Parkinson’s disease: Who gets it early? Parkinsonism & Related Disorders 2017; 44: 91–94.

[46] Postuma RB, Berg D, Stern M, et al. MDS clinical diagnostic criteria for Parkinson’s disease. Mov Disord 2015; 30: 1591–1601.

[47] Schade S, Mollenhauer B, Trenkwalder C. Levodopa Equivalent Dose Conversion Factors: An Updated Proposal Including Opicapone and Safinamide. Movement Disorders Clinical Practice 2020; 7: 343–345.

[48] Hoehn MM, Yahr MD. Parkinsonism: onset, progression and mortality. Neurology 1967; 17: 427–442.

[49] Goetz CG, Tilley BC, Shaftman SR, et al. Movement Disorder Society-sponsored revision of the Unified Parkinson’s Disease Rating Scale (MDS-UPDRS): scale presentation and clinimetric testing results. Mov Disord 2008; 23: 2129–2170.

[50] Zigmond AS, Snaith RP. The hospital anxiety and depression scale. Acta Psychiatr Scand 1983; 67: 361–370.

[51] Nasreddine ZS, Patel BB. Validation of Montreal Cognitive Assessment, MoCA, Alternate French Versions. Can J Neurol Sci 2016; 43: 665–671.

[52] Reitan RM, Wolfson D. The Halstead-Reitan Neuropsychological Test Battery: Theory and Clinical Interpretation. Neuropsychology Press, 1985.

[53] Stroop JR. Studies of interference in serial verbal reactions. J Exp Psychol 1935; 643–662.

[54] Delis DC, Kaplan E, Kramer JH. Delis-Kaplan Executive Function System. Epub ahead of print 8 October 2012. DOI: 10.1037/t15082-000.

[55] Blanke O, Bernasconi F, Potheegadoo J. Phantom Boarder Relates to Experimentally-Induced Presence Hallucinations in Parkinson’s Disease. Mov Disord Clin Pract 2023; 10: 617–624.

[56] Gryc W, Roberts KA, Zabetian CP, et al. Hallucinations and Development of Dementia in Parkinson’s Disease. J Parkinsons Dis 2020; 10: 1643–1648.

[57] Flanigan JL, Harrison MB, Patrie JT, et al. Clinical and cognitive features associated with psychosis in Parkinson’s disease: a longitudinal study. Front Aging Neurosci 2024; 16: 1463426.

[58] Ozawa M, Shiraishi T, Murakami H, et al. Structural MRI study of Pareidolia and Visual Hallucinations in Drug-Naïve Parkinson’s disease. Sci Rep 2024; 14: 31293.

[59] Stam CJ, Visser SL, Op de Coul AA, et al. Disturbed frontal regulation of attention in Parkinson’s disease. Brain 1993; 116 ( Pt 5): 1139–1158.

[60] Costa A, Peppe A, Mazzù I, et al. Dopamine Treatment and Cognitive Functioning in Individuals with Parkinson’s Disease: The “Cognitive Flexibility” Hypothesis Seems to Work. Behavioural Neurology 2014; 2014: e260896.

[61] Zarkali A, Luppi AI, Stamatakis EA, et al. Changes in dynamic transitions between integrated and segregated states underlie visual hallucinations in Parkinson’s disease. Commun Biol 2022; 5: 928.

[62] Dujardin K, Duhamel A, Delliaux M, et al. Cognitive complaints in Parkinson’s disease: its relationship with objective cognitive decline. J Neurol 2010; 257: 79–84.

[63] Pozeg P, Rognini G, Salomon R, et al. Crossing the hands increases illusory self-touch. PLoS ONE 2014; 9: e94008.

[64] Weiskrantz L, Elliott J, Darlington C. Preliminary observations on tickling oneself. Nature 1971; 230: 598–599.

[65] Blakemore SJ, Smith J, Steel R, et al. The perception of self-produced sensory stimuli in patients with auditory hallucinations and passivity experiences: evidence for a breakdown in self-monitoring. Psychol Med 2000; 30: 1131–1139.

[66] Schneider RB, Mills KA, Nirenberg MJ, et al. Defining the Importance of Minor Hallucinations in Parkinson’s Disease. Movement Disorders 2024; 39: 1083–1083.

[67] Aarsland D, Batzu L, Halliday GM, et al. Parkinson disease-associated cognitive impairment. Nat Rev Dis Primers 2021; 7: 1–21.

[68] Fénelon G, Alves G. Epidemiology of psychosis in Parkinson’s disease. J Neurol Sci 2010; 289: 12–17.

[69] Omoto S, Murakami H, Shiraishi T, et al. Risk factors for minor hallucinations in Parkinson’s disease. Acta Neurol Scand 2021; 143: 538–544.

[70] Zhong M, Gu R, Zhu S, et al. Prevalence and Risk Factors for Minor Hallucinations in Patients with Parkinson’s Disease. Behav Neurol 2021; 2021: 3469706.

[71] Forsaa EB, Larsen JP, Wentzel-Larsen T, et al. A 12-Year Population-Based Study of Psychosis in Parkinson Disease. Archives of Neurology 2010; 67: 996–1001.

[72] Goetz CG, Leurgans S, Pappert EJ, et al. Prospective longitudinal assessment of hallucinations in Parkinson’s disease. Neurology 2001; 57: 2078–2082.

[73] Zhu K, Hilten JJ van, Putter H, et al. Risk factors for hallucinations in Parkinson’s disease: Results from a large prospective cohort study. Movement Disorders 2013; 28: 755–762.

[74] Crook T, Ferris S, Bartus R. Assessment in Geriatric Psychopharmacology. Mark Powley Associates, 1983.

[75] Ko J, Ha J, Lee JJ, et al. Reliability and Validity of the Subjective Cognitive Complaints Questionnaire for Parkinson’s Disease (SCCQ-PD). Journal of Clinical Neurology 2022; 18: 171–178.

