## Supplementary material for "Frontal subcortical executive dysfunction and minor hallucinations in Parkinson’s disease are linked to sensitivity to somatomotor conflicts"

**Affiliation:**

Laboratory of Cognitive Neuroscience

Bertarelli Foundation Chair in Cognitive Neuroprosthetics

Neuro-X Institute, Campus Biotech

Chemin des Mines 9,

1202 Geneva

Switzerland

+41 21 693 96 21

Dr. Jevita Potheegadoo

Laboratory of Cognitive Neuroscience

Neuro-X Institute, Campus Biotech,

Chemin des Mines 9,

1202 Geneva

Switzerland

+41 21 693 95 68

### Supplemental Material 1: Methods

#### Neuropsychological assessment

The Stroop test [1] used in the current study included four conditions: color naming (condition 1), color reading (condition 2), an inhibitory control/interference condition during which participants name the color of the ink rather than the word itself (condition 3), and a condition (condition 4) measuring cognitive flexibility. In the latter condition, two types of stimuli are presented: participants name the ink color of the word, but if a colored word is framed within a box, they instead read the word itself.

### Supplemental Material 2: Types and co-occurrence of MH among the PD-MH group

In the PD-MH group, passage hallucinations were the most common (61.29%), followed by visual illusions/misperceptions (58.06%) and presence hallucinations (54.84%) (**Figure S1**). Six patients experienced isolated passage hallucinations, 4 patients reported isolated PH and 7 had visual illusions/misperceptions only. Nine patients experienced the 3 types of MH, 3 had passage hallucinations and PH, and 1 patient had both passage hallucinations and visual illusions/misperceptions, and 1 has a combination of PH and visual illusions/misperceptions.

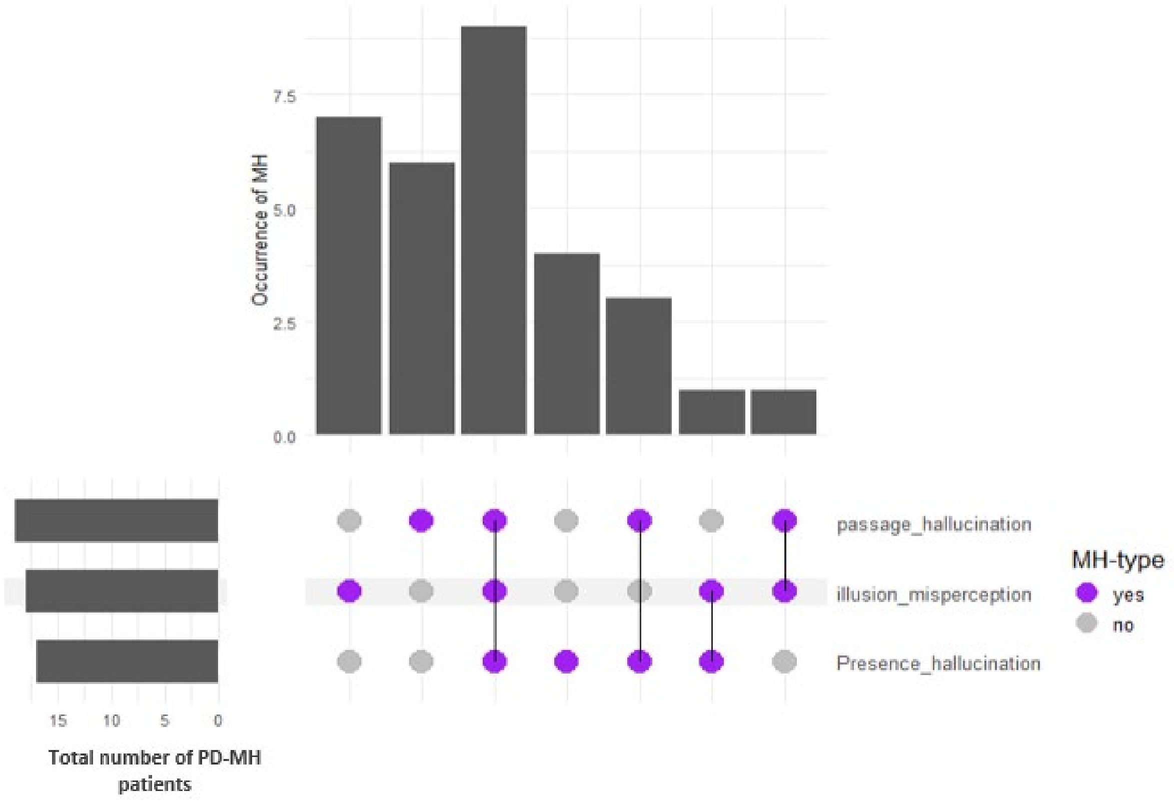

**Figure S1. Distribution and overlap of minor hallucination (MH) types in in the PD-MH group.** The UpSet plot illustrates the frequency and co-occurrence of different MH types across patients. Vertical bars represent the number of participants exhibiting each specific combination of MH types, with higher bars indicating more frequent occurrences of that combination. Dots and connecting lines in the matrix indicate which MH types are present (purple dots) or absent (grey dots) in each intersection. Bars are ordered by relative frequency (ratio), and the left annotation shows the overall occurrence of MH.

### Supplemental Material 3: Relationship between riPH and neuropsychology

PD-CRS

To complement the main analyses, we examined the other main effects and interactions of frontal subcortical cognitive scores (from the PD-CRS), subgroup, and delay on riPH sensitivity. A main effect of group showed a trend toward significance (β=1.31, SE=0.69, z=1.88, *P*= 0.059), suggesting a tendency for the PD-MH group to present higher riPH sensitivity than the PD-nH one. A significant main effect of frontal subcortical cognitive scores (β=−1.39, SE=0.70, z=−1.98, *P*=0.048) indicated that lower frontal subcortical scores were associated with higher riPH sensitivity. A significant main effect of delay (β=0.64, SE=0.10, z=6.73, *P*< 0.001) indicated that riPH sensitivity increased with longer delays of somatomotor conflicts. The two-way interactions between group and frontal subcortical scores (β=−0.47, SE=0.73, z=−0.64, *P*=0.52) and between group and delay (β=0.05, SE=0.10, z=0.54, *P*=0.59) were not significant, suggesting that the effects of cognitive performance and delay did not differ between PD-MH and PD-nH groups. The interaction between frontal subcortical scores and delay was not significant (β=−0.18, SE=0.10, z=−1.72, *P*=0.09).

Furthermore, our analysis of posterior cortical scores (PD-CRS) showed a significant main effect of group (β=1.82, SE=0.70, z=2.59, *P*=0.01), with PD-MH patients showing higher riPH sensitivity than PD-nH patients, and a significant main effect of delay (β=0.71, SE=0.10, z=6.88, *P*<0.001) with riPH sensitivity increasing at longer delays. Posterior cortical scores showed no significant main effect (β=1.02, SE=0.69, z=1.49, *P*=0.14). No significant two-way interactions were found between group and posterior cortical scores (β=−0.01, SE=0.70, z=−0.02, *P*=0.99), between group and delay (β=−0.05, SE=0.10, z=−0.52, *P*=0.60) or between posterior cortical scores and delay (β=−0.21, SE=0.17, z=−1.19, *P*=0.23). Post-hoc group-specific analyses indicated no significant effects of posterior cortical scores on riPH sensitivity in either group. In PD-nH patients, the interaction between posterior cortical scores and delay was not significant (β=−0.56, SE=0.33, z=−1.73, $P$=0.08). Overall, posterior cortical cognitive performance alone does not appear to modulate riPH sensitivity in either group.

### Stroop color-word test

#### Condition 3

Examining the number of self-corrected errors at the Stroop (inhibitory control, condition 3), there was a significant main effect of group on riPH sensitivity (β=1.90, SE=0.69, z=2.73, $P$=0.01), with PD-MH patients showing higher riPH sensitivity than PD-nH patients, and a significant main effect of delay (β=0.64, SE=0.10, z=6.22, $P$< 0.001), with riPH sensitivity increasing at longer delays. No significant main effect of Stroop self-corrected error rate was observed (β=−0.27, SE=0.85, z=−0.32, $P$=0.75), nor were any two-way interactions significant. The three-way interaction between group, Stroop self-corrected error rate, and delay approached trend-level significance (β=0.28, SE=0.15, z=1.88, $P$=0.06).

Analyses on the number of non-corrected errors showed a significant main effect of group on riPH sensitivity (β=2.23, SE=0.75, z=2.99, $P$=0.003), with PD-MH patients showing higher riPH sensitivity than PD-nH patients, and a significant main effect of delay (β=0.97, SE=0.17, z=5.85, $P$<0.001) with riPH sensitivity increasing with longer delays. Stroop non-corrected error rate showed no significant main effect on riPH sensitivity (β=−1.43, SE=1.01, z=−1.42, $P$=0.16). A significant three-way interaction between group, the number of non-corrected errors, and delay (β=−1.22, SE=0.49, z=−2.51, $P$=0.01) indicated that the relationship between Stroop error rate and riPH sensitivity differed across groups and levels of delay asynchrony. In addition, a significant interaction between group and Stroop error rate was observed (β=1.99, SE=1.01, z=1.97, $P$=0.049), indicating that the association between executive performance and riPH sensitivity differed between PD-MH and PD-nH patients.

However, follow-up group-specific analyses did not show a significant main effect of the Stroop error rate on riPH sensitivity in the PD-MH group (β=0.49, SE=0.68, z=0.72, $P$=0.47), indicating that executive performance did not modulate riPH sensitivity in this group. In the PD-nH group, as Stroop non-corrected errors increases, the effect of delay on riPH sensitivity increased also as indicated by a significant interaction between Stroop error rate and delay ($\beta$=2.68, SE=0.99, z=2.72, *P*=0.007).

Together, these findings indicate that executive performance, as measured by Stroop non-corrected errors, modulated the effect of delay on riPH sensitivity in PD-nH patients but not in PD-MH patients, reflecting group-specific differences in the relationship between inhibitory control and susceptibility to riPH.

Condition 4

Analyses of number of self- and non-corrected errors at the Stroop test, condition 4 (mental flexibility) showed significant main effects of group on riPH sensitivity (self-corrected: $\beta$=2.01, SE=0.73, z=2.75, *P*=0.01; non-corrected: $\beta$=1.96, SE=0.72, z=2.73, *P*=0.01), with the PD-MH patients being more sensitive to riPH, and delay (self-corrected: $\beta$=0.65, SE=0.10, z=6.59, *P*<0.001; non-corrected: $\beta$=0.62, SE=0.10, z=6.25, *P*<0.001), showing increase in riPH sensitivity at longer delays. Neither self-corrected nor non-corrected Stroop errors had significant main effects (all *P*>0.50), and no two-way or three-way interactions with group or delay were significant (all *P*>0.34). These results indicate that mental flexibility, as measured by the number of errors at the Stroop test (condition 4), did not modulate riPH sensitivity in either group.

Trail Making Test (TMT)

TMT (part A)

Analyses of TMT part A (self-corrected and non-corrected errors) showed no significant main effects or interactions with group or delay on riPH sensitivity (all *P*>0.35). Convergence issues were noted for the self-corrected errors model, but parameter estimates indicated no meaningful effects. These results suggest that performance on TMT-A did not modulate riPH sensitivity in either PD-MH or PD-nH patients.

TMT (part B)

Analyses of TMT part B (self-corrected and non-corrected errors) showed a significant main effect of group on riPH sensitivity (self-corrected: β=1.78, SE=0.73, z=2.43, *P*=0.02; non-corrected: β=1.75, SE=0.70, z=2.49, *P*=0.01) with PD-MH patients being more sensitive to riPH, and a significant main effect of delay (self-corrected: β=0.66, SE=0.10, z=6.48, *P*<0.001; non-corrected: β=0.69, SE=0.10, z=7.02, *P*<0.001) showing increased riPH sensitivity at longer delays.

For self-corrected errors, a significant three-way interaction between group, error rate, and delay was observed (β=−0.20, SE=0.08, z=−2.59, *P*=0.01), suggesting that the influence of s errors on riPH sensitivity across delays differed between groups. Post-hoc group-specific analyses revealed no significant main or interaction effects in either PD-MH (interaction between self-corrected error rate and delay: β=−0.16, SE=0.09, z=−1.67, *P*=0.09) or PD-nH patients (interaction between self-corrected error rate and delay: β=0.24, SE=0.12, z=1.91, *P*=0.06), indicating that performance at TMT part B did not directly modulate riPH sensitivity within each group.

Non-corrected errors showed a similar pattern, with no significant interactions or main effects beyond the main effects of group and delay (all *P*>0.18).

### Supplemental References

[1] Delis DC, Kaplan E, Kramer JH. Delis-Kaplan Executive Function System. Epub ahead of print 8 October 2012. DOI: 10.1037/t15082-000.

[2] Pagonabarraga J, Kulisevsky J, Llebaria G, et al. Parkinson's disease-cognitive rating scale: a new cognitive scale specific for Parkinson's disease. *Mov Disord* 2008; 23: 998–1005.

[3] Army Individual Test Battery. *Manual of directions and scoring*. Washington, DC: War Department, Adjunct General's Office, 1944.

[4] Goetz CG, Tilley BC, Shaftman SR, et al. Movement Disorder Society-sponsored revision of the Unified Parkinson's Disease Rating Scale (MDS-UPDRS): scale presentation and clinimetric testing results. *Mov Disord* 2008; 23: 2129–2170.
